# Research transparency promotion by surgical journals publishing randomised controlled trials: a survey

**DOI:** 10.1101/19002386

**Authors:** N. Lombard, A. Gasmi, L. Sulpice, K. Boudjema, F. Naudet, D. Bergeat

## Abstract

**Objective:** To describe the surgical journal position statement on data-sharing policies (primary objective) and to describe the other features of their research transparency promotion.

**Methods:** Only “SURGICAL” journals with an impact factor superior to 2 (Web of Science) were eligible for the study. They were not included if there were no explicit instructions for clinical trial publication in the instructions for authors and if there were no RCT published between January 2016 and January 2019. The primary outcome was the existence of a data-sharing policy in the instructions for authors. Details on research transparency promotion were also collected, namely the existence of a “prospective registration of clinical trials requirement” policy; a “COIs” disclosure requirement and a specific reference to reporting guidelines such as CONSORT for RCT.

**Results:** Among the 87 surgical journals eligible, 82 (94%) were included in the analysis: 67 (77%) had explicit instructions for RCT and of the remaining, 15 (17.2%) had published at least one RCT between 2016-2019. The median impact factor was 2.98 [IQR=2.48-3.77] and in 2016 and 2017, the journals published a median of 11.5 RCT [IQR=5-20.75]. Data-sharing statement instructions (primary outcome) were ICMJE-compliant in four cases (4.88%), weaker in 45.12% (n=37) and inexistent in 50% (n=41) of the journals. As for data-sharing statements, no association was found between journal characteristics and the existence of data-sharing policies (ICMJE-compliant or weaker). A “prospective registration of clinical trials requirement” was associated with ICMJE allusion or affiliation and higher impact factors. Journals with specific RCT instructions in their OIA and journals referenced on the ICMJE website more frequently mandated the use of CONSORT guidelines.

**Conclusion:** Research transparency promotion is still limited in surgical journals. Uniformization of journal requirements vis-à-vis ICMJE guidelines could be a first step forward for research transparency promotion in surgery.

## INTRODUCTION

Surgical journals have a key role to ensure transparency, openness, and reproducibility^1^ – features that are expected to increase value and reduce waste in the research they publish^2^. The highest editorial standards are expected when it comes to randomised controlled trials (RCT) because their importance is paramount in drafting guidelines that can impact medical practice worldwide. The latest breakthrough was the adoption by the International Committee of Medical Journal Editors (ICMJE) of a policy that encourages RCT data sharing and requires a data-sharing statement to be included in the reports of published clinical trials^3^. Other aspects of research transparency promotion have been previously promoted such as registration of the trial^4^, adoption of the CONSORT statement^5^ and declaration of conflicting interests (COI)^6^. However, transparent practices in the surgical community could be suboptimal as suggested by the underreporting of COI^7^.

The aim of this study is to describe the surgical journal position statement on data-sharing policies (primary objective) and to describe the other features of their research transparency promotion.

## METHODS

This survey of surgical journals was registered with a protocol in the Open Science Framework on February, 25^th^ 2019.

(https://osf.io/d6bua/?view_only=6d0a6290df804f8a843ad8cddead81e5)

### Eligibility criteria and Journal selection

Two reviewers (NL and AG) used Web of Science to select journals classified in the “SURGICAL” category with an impact factor superior to 2. Surgical journals were not included if there were no explicit instructions for clinical trial publication in the instructions for authors and if there were no RCT published between January 2016 and January 2019. Two authors (NL and AG) independently extracted the data. Disagreements were resolved by consensus or in consultation with a third reviewer (DB). The list of journals was extracted in December 2018 and the official instructions for authors (OIA) were downloaded on January 13-14, 2019.

Our primary outcome was the existence of a data-sharing policy in the instructions for authors. Types of policies for data sharing were described using the following classification: “ICMJE compliant” (policies explicitly stating that the data-sharing statement was mandatory), “Weaker policy” (policies stating a data-sharing statement could be included in the paper), “None” (no mention of any data-sharing policy). Details on research transparency promotion were also collected, namely the existence of a “prospective registration of clinical trials requirement” policy; a “COIs” disclosure requirement and a specific reference to reporting guidelines such as CONSORT for RCT. Various journal features were also extracted (see Table 1). Journal impact factors were extracted from the Web of Science data base and the number of RCT published between January 2016 and January 2018 was extracted from Pubmed (we initially planned to evaluate the number of RCT published in 2016, 2017 and 2018 but this was not possible because at the time of data extraction, all RCT published were not fully indexed in Pubmed). ICMJE “affiliation” was defined as journals referenced as “Journals stating that they follow ICMJE Recommendations” at: http://www.icmje.org/journals-following-the-icmje-recommendations.

**Table 1:** Journal characteristics and policies. OIA = official instructions for authors, ICMJE = International Committee of Medical Journal Editors.

|  |  |  |
| --- | --- | --- |
| **Explicit instruction(s) for RCT in the OIA** |  |  |
|  | Yes | 67 (81.71) |
|  | No | 15 (18.29) |
| **Allude to ICMJE guidelines in the OIA** |  |  |
|  | Yes | 63 (76.83) |
|  | No | 19 (23.17) |
| **ICMJE affiliation \*** |  |  |
|  | Yes | 35 (42.68) |
|  | No | 47 (57.32) |
| **Data sharing statement policies** |  |  |
|  | ICMJE compliant | 4 (4.88) |
|  | Weaker | 37 (45.12) |
|  | None | 41 (50.00) |
| **Prospective registration of clinical study policies** |  |  |
|  | Mandatory | 53 (64.63) |
|  | Weaker | 29 (35.37) |
| **Consort guideline policies** |  |  |
|  | Mandatory | 24 (29.27) |
|  | Weaker | 58 (70.73) |
| **COI disclosure policies** |  |  |
|  | Mandatory | 77 (93.90) |
|  | Weaker | 5 (6.10) |
| **Publisher** |  |  |
|  | Elsevier | 25 (30.49) |
|  | Springer Nature | 13 (15.85) |
|  | Wiley Online Library | 12 (14.63) |
|  | Wolters Kluwer | 11 (13.41) |
|  | Other | 21 (25.61) |
| **Geographical area of major editorial committee** |  |  |
|  | Asia | 6 (7.32) |
|  | Europe | 26 (31.71) |
|  | International | 4 (4.88) |
|  | Northern America | 46 (56.10) |
| **Journal topics** |  |  |
|  | Generalist | 20 (24.39) |
|  | Specialist | 62 (75.61) |
| **Publishing model** |  |  |
|  | Hybrid | 73 (89.02) |
|  | Open Access | 1 (1.22) |
|  | Paywall | 8 (9.76) |
| **Number of RCT published in 2016-2017** |  | 11.5 [5-20.75] |
| **Journal impact factor** |  | 2.98 [2.48-3.77] |

Quantitative variables are reported with number and (%); Quantitative variables are reported with median and [Interval inter quartil range (IQR)]; OIA= official instructions for authors ; \* ICMJE afiliation = Journals referenced as "Journals stating that they follow the ICMJE Recommendations" on http://www.icmje.org/journals-following-the-icmje-recommendations/

### Statistical analyses

Analyses of all included journals were performed using open source R statistical software (http://www.r-project.org/). Quantitative variables were expressed with median and the interquartile range (IQR) and compared with the Mann-Whitney U test. Qualitative variables were expressed as a percentage and compared with a Chi-squared test or Fisher test as appropriate. Univariate exploratory analyses were performed to explore the associations between journal features and the various transparency policies. Multivariate analyses were planned but not run owing to sparse data.

## RESULTS

Among the 87 surgical journals eligible, 82 (94%) were included in the analysis: 67 (77%) had explicit instructions for RCT and of the remaining, 15 (17.2%) had published at least one RCT between 2016-2019 (**Supp. figure 1** details the selection process). The characteristics of these journals are detailed in **Table1**. The median impact factor was 2.98 [IQR=2.48-3.77]. In 2016 and 2017, the journals published a median of 11.5 RCT [IQR=5-20.75]. The publishing model was “hybrid” in most cases (89.02%) and North America was the principal geographical area of journal editorial committees (56.10%) Data-sharing statement instructions were ICMJE-compliant in four cases (4.88%), weaker in 45.12% (n=37) and inexistent in 50% (n=41) of the journals. COI disclosure was mandatory in 77 journals (93.90%). A reference to CONSORT guidelines was made in 24 journals (29.27%). Prospective registration of clinical trials was mandatory in 53 cases (64.63%). **Figure 1** presents the relationship between the different research transparency promotion items and journal impact factors and the number of RCT published between 2016 and 2018. The associations between journal features and the different transparency policies are presented in **Supp. Table 1**. As for data-sharing statements, no association was found between journal characteristics and the existence of data-sharing policies (ICMJE-compliant or weaker). A “prospective registration of clinical trials requirement” was associated with ICMJE allusion or affiliation and higher impact factors. Journals with specific RCT instructions in their OIA and journals referenced on the ICMJE website more frequently mandated the use of CONSORT guidelines. No other association was found.

**Figure 1:**
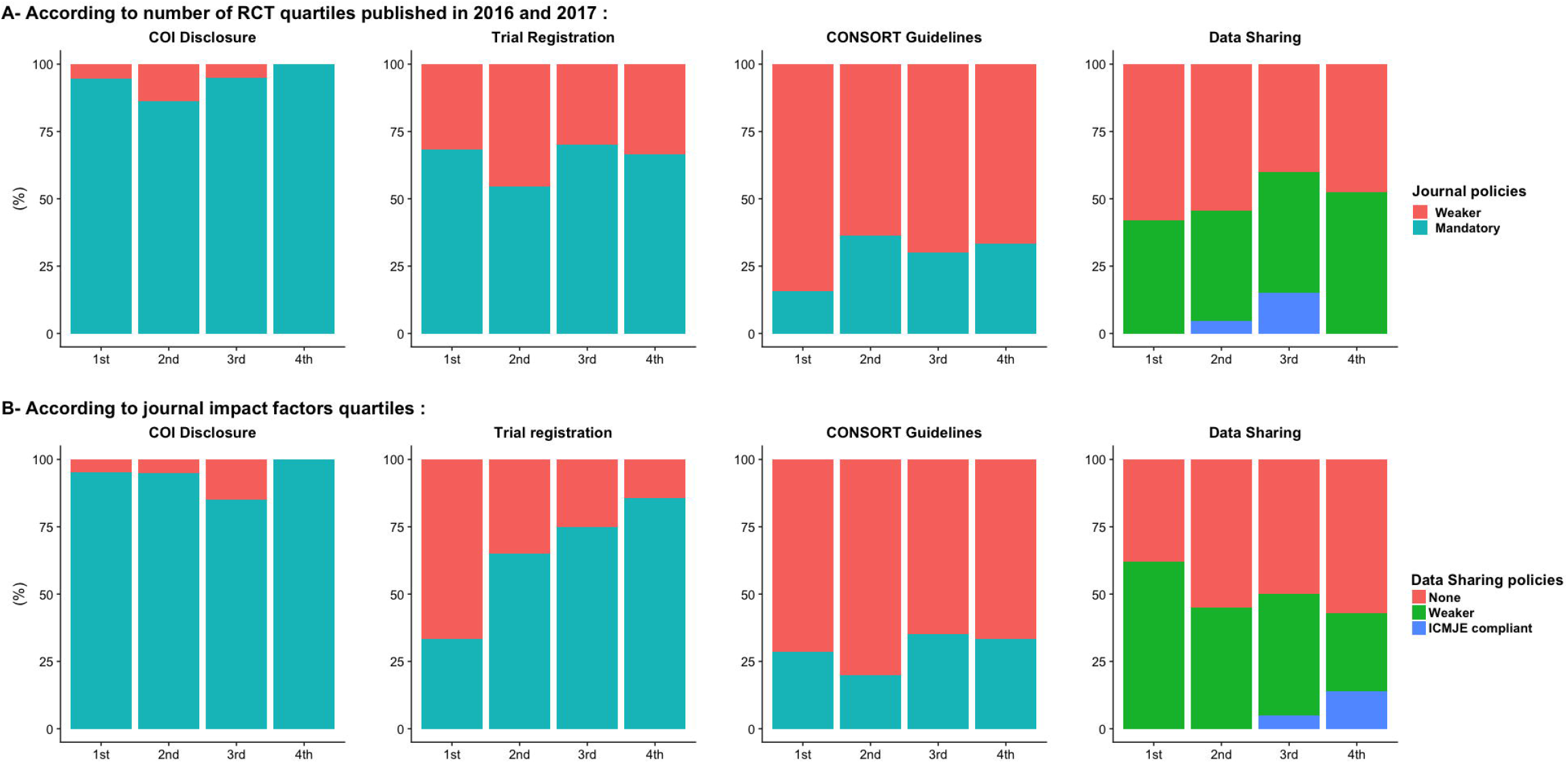
Research transparency promotion related A) to the number of RCT quartiles published in 2016 and 2017 and (B) to journal impact factor quartiles. COI = Conflict of interest; RCT = randomised controlled trial.

## DISCUSSION

We noted low rates of implementation of data-sharing policies, i.e. 50 % of the journals had no explicit policy included in their instructions for authors. When explicit, these policies were rather allusive and weaker than the ICMJE recommendation that make a data-sharing statement mandatory for RCT. Of course, we studied a moving target and one could argue that the ICMJE position on data sharing was fairly recent (data extracted 6 months after the ICMJE statement) and that a number of journals did not have the time to implement it when our survey was conducted. However, this policy was announced in 2017^8^ and 35 (43%) journals are listed on the ICMJE website. Interestingly, implementation of older policies was also suboptimal, even for making a specific reference to reporting guidelines such as CONSORT for RCT which date from 1996^5^. Except for COI disclosure, those policies were mostly non-binding. These disappointing results are not new. In 2014, Chapman et al.^9^ warned about sub-optimal transparency policies in 10 leading surgical journals.

We considered a journal’s policies presented on its website as a surrogate marker of implementation of these policies. However, it is possible that editors of journals with a policy do not implement them in an optimal manner^10^ or, conversely, that a journal with no specific policy documented on the website requires authors to comply with some of the features we explored. Of note, previous research has shown that journal requirements can have a significant impact on changing researcher practices^11^ but an obvious next step is to explore the transparency features of the published RCT in these journals.

Of concern, we found no association of research transparency items with impact factors nor with the number of RCT published except for prospective trial registration among the surveyed surgical journals. This is of concern since impact factor (rounded to the nearest two decimals) is misused as a surrogate to assess the quality of a given journal and sometimes of an individual paper.^12,13^

## CONCLUSION

As part of a wider movement^14^, we suggest that indicators of quality such as prospective audits of policies and published papers must be used to assess journals instead of journal impact factors. We encourage surgical journals to be part of the move to improve their research transparency promotion. Uniformization of journal requirements vis-à-vis ICMJE guidelines could be a first step forward for research transparency promotion in surgery.

## Data Availability

supporting data and analyze codes are available on open science frame work

## List of abbreviations

RCT: Randomised Controlled Trial
ICMJE: International Committee of Medical Journal Editors
COI: Conflict of Interest
OIA: Official Instructions for Authors
IQR: Interquartile interval range

## Author contributions

Study concept and design: DB, FN

Acquisition of data: NL, AG, DB

Analysis and interpretation of data: DB, NL, FN

Drafting of the manuscript: DB, NL, FN

Critical revision of the manuscript for important intellectual content: FN, KB, LS, AG

Statistical analysis: DB, FN

## Acknowledgements

Study protocol is already available on Open Science Framework. Data extracted and statistical code is available on Open Science Framework.

## Compliance with Ethical Standards

### Funding

There was no financial support for this study. Florian Naudet’s works on reproducibility are part of the ReiTheR (reproducibility in therapeutic research) project. This project is funded by the French National Research Agency (ANR, reference number ANR-17-CE-36-0010-01). The sponsor had no role concerning preparation, review, or approval of the manuscript.

### Ethical approval

For this type of study, formal consent is not required.

## Table titles and legends

**Supplementary Table 1:** Exploratory analysis, association between journal features and transparency policies with univariate analysis. OIA = official instructions for authors, ICMJE = International Committee of Medical Journal Editors.

## Figure titles and legends

**Supplementary Figure 1:** Journal selection process. RCT = randomised controlled trial.

## REFERENCES

1 Nosek BA, Alter G, Banks GC, Borsboom D, Bowman SD, Breckler SJ, et al. Promoting an open research culture. Science. 2015 Jun 26; 348: 1422–1425.

2 Macleod MR, Michie S, Roberts I, Dirnagl U, Chalmers I, Ioannidis JPA, et al. Biomedical research: increasing value, reducing waste. The Lancet. 2014 Jan; 383: 101–104.

3 Taichman DB, Sahni P, Pinborg A, Peiperl L, Laine C, James A, et al. Data Sharing Statements for Clinical Trials - A Requirement of the International Committee of Medical Journal Editors. N Engl J Med. 2017 08; 376: 2277–2279.

4 De Angelis C, Drazen JM, Frizelle FA, Haug C, Hoey J, Horton R, et al. Clinical trial registration: a statement from the International Committee of Medical Journal Editors. N Engl J Med. 2004 Sep 16; 351: 1250–1251.

5 Limb C, White A, Fielding A, Lunt A, Borrelli MR, Alsafi Z, et al. Compliance of Randomized Controlled Trials Published in General Surgical Journals With the CONSORT 2010 Statement. Ann Surg. 2019 Mar; 269: e25–e27.

6 Conflict of interest. International Committee of Medical Journal Editors. Ann Intern Med. 1993 Apr 15; 118: 646–647.

7 Ziai K, Pigazzi A, Smith BR, Nouri-Nikbakht R, Nepomuceno H, Carmichael JC, et al. Association of Compensation From the Surgical and Medical Device Industry to Physicians and Self-declared Conflict of Interest. JAMA Surg. 2018 Aug 15;

8 Taichman DB, Sahni P, Pinborg A, Peiperl L, Laine C, James A, et al. Data Sharing Statements for Clinical Trials: A Requirement of the International Committee of Medical Journal Editors. Ann Intern Med. 2017 04; 167: 63–65.

9 Promoting transparency in clinical research: Systematic review of disclosure and data-sharing policies in surgical journals | S.J. Chapman | Request PDF [Internet]. ResearchGate. [cited 2019 Jun 26]. Available from: https://www.researchgate.net/publication/275535694_Promoting_transparency_in_clinical_research_Systematic_review_of_disclosure_and_data-sharing_policies_in_surgical_journals

10 Naudet F, Sakarovitch C, Janiaud P, Cristea I, Fanelli D, Moher D, et al. Data sharing and reanalysis of randomized controlled trials in leading biomedical journals with a full data sharing policy: survey of studies published in The BMJ and PLOS Medicine. BMJ. 2018 Feb 13; k400.

11 Giofrè D, Cumming G, Fresc L, Boedker I, Tressoldi P. The influence of journal submission guidelines on authors’ reporting of statistics and use of open research practices. PloS One. 2017; 12: e0175583.

12 Ioannidis JPA, Thombs BD. A user’s guide to inflated and manipulated impact factors. Eur J Clin Invest. 0: e13151.

13 San-Francisco-Declaration-on-Research-Assessment.pdf [Internet]. [cited 2019 Jun 26]. Available from: https://www.ouvrirlascience.fr/wp-content/uploads/2019/03/San-Francisco-Declaration-on-Research-Assessment.pdf

14 Moher D, Naudet F, Cristea IA, Miedema F, Ioannidis JPA, Goodman SN. Assessing scientists for hiring, promotion, and tenure. PLOS Biol. 2018 Mar 29; 16: e2004089.

